# An algorithm to build synthetic temporal contact networks based on close-proximity interactions data

**DOI:** 10.1101/2023.10.23.23296945

**Authors:** Audrey Duval, Quentin Leclerc, Didier Guillemot, Laura Temime, Lulla Opatowski

**Affiliations:** Institut Pasteur, Université Paris Cité, Epidemiology and Modelling of Bacterial Escape to Antimicrobials (EMEA), 75015 Paris, France; INSERM, Université Paris-Saclay, Université de Versailles St-Quentin-en-Yvelines, Team Echappement aux Anti-infectieux et Pharmacoépidémiologie U1018, CESP, 78000 Versailles, France; Laboratoire Modélisation, Epidémiologie et Surveillance des Risques Sanitaires (MESuRS), Conservatoire National des Arts et Métiers, 73003 Paris, France; AP-HP, Paris Saclay, Department of Public Health, Medical Information, Clinical research, F-92380, Garches; Institut Pasteur, Conservatoire National des Arts et Métiers, Unité PACRI, 75015 Paris, France

**Keywords:** long-term care facility, contact network, close-proximity interactions, sensors, network reconstruction

## Abstract

Small populations (e.g., hospitals, schools or workplaces) are characterised by high contact heterogeneity and stochasticity affecting pathogen transmission dynamics. Empirical individual contact data provide unprecedented information to characterize such heterogeneity and are increasingly available, but are usually collected over a limited period, and can suffer from observation bias. We propose an algorithm to stochastically reconstruct realistic temporal networks from individual contact data in healthcare settings (HCS) and test this approach using real data previously collected in a long-term care facility (LTCF).

Our algorithm generates full networks from recorded close-proximity interactions, using hourly inter-individual contact rates and information on individuals’ wards, the categories of staff involved in contacts, and the frequency of recurring contacts. It also provides data augmentation by reconstructing contacts for days when some individuals are present in the HCS without having contacts recorded in the empirical data. Recording bias is formalized through an observation model, to allow direct comparison between the augmented and observed networks. We validate our algorithm using data collected during the i-Bird study, and compare the empirical and reconstructed networks.

The algorithm was substantially more accurate to reproduce network characteristics than random graphs. The reconstructed networks reproduced well the assortativity by ward (first– third quartiles observed: 0.54–0.64; synthetic: 0.52–0.64) and the hourly staff and patient contact patterns. Importantly, the observed temporal correlation was also well reproduced (0.39–0.50 vs 0.37–0.44), indicating that our algorithm could recreate a realistic temporal structure. The algorithm consistently recreated unobserved contacts to generate full reconstructed networks for the LTCF.

To conclude, we propose an approach to generate realistic temporal contact networks and reconstruct unobserved contacts from summary statistics computed using individual-level interaction networks. This could be applied and extended to generate contact networks to other HCS using limited empirical data, to subsequently inform individual-based epidemic models.

**Author summary:** Contact networks are the most informative representation of the contact heterogeneity, and therefore infectious disease transmission risk, in small populations. However, the data collection required is costly and complex, usually limited to a few days only and likely to suffer from partially observed data, making the practical integration of networks into models challenging. In this article, we present an approach leveraging empirical individual contact data to stochastically reconstruct realistic temporal networks in healthcare settings. The algorithm accounts for population specificities including the hourly distribution of contact rates between different individuals (staff categories, patients) and the probability for contact repetition between the same individuals. We illustrate and validate this algorithm using a real contact network measured in a long-term care facility. Our approach outperforms random graphs informed by the same data to accurately reproduce observed network characteristics and hourly staff-patient contact patterns. The algorithm recreates unobserved contacts, providing data augmentation for times with missing information. This method should improve the usability and reliability of contact networks, and therefore promote integration of empiric contact data in individual-based models.

## Introduction

Limiting the burden of infectious diseases requires a good understanding of how they spread. For pathogens transmitted mostly via close-proximity interactions, the rate at which individuals come into contact with each other is strongly correlated with the expected spread of the disease across the population [1]. In large populations such as cities or countries, contact structures are usually approximated by grouping individuals into relatively broad categories (neighbourhood, age…), and assuming that contact rates are heterogeneous between categories, but homogeneous within [2,3]. In small populations such as healthcare institutions, schools, or workplaces however, disease transmission is affected by high contact heterogeneity and stochasticity [4]. Capturing these characteristics requires a detailed, individual-level description of contacts instead of only relying on summary contact rates by groups [5,6].

Contact networks are increasingly used to fully capture the interactions between individuals in small populations [7,8]. These networks explicitly represent the links between all individuals in such populations, as opposed to contact matrices which capture average contact rates between groups of individuals [9,10]. Temporal contact networks further capture the time-changing nature of contacts, therefore representing individual interactions more accurately than static networks [11–15]. Contact networks can be coupled with individual-based mathematical models to help design effective interventions against the spread of infectious diseases, since they enable the identification of highly connected individuals who can be targeted to lead to the greatest impact on transmission [10]. Recently, empirical data collected to build inter-individual temporal networks has become increasingly available to inform contact networks. For example, studies have used sensors to record close-proximity interactions between individuals [16–18], and contact tracing programs have relied on the integrated Bluetooth technology in mobile phones [19].

However, the detailed empirical data required to build temporal contact networks remain subject to several limitations [20,21]. These data are typically collected over a few days only [22,23], and may be subject to observation bias; sensors might not be properly placed to register contacts [24], or individuals may disable Bluetooth on their mobile phones at different times [19]. Due to the resulting missed contacts, the networks derived from these data may only be partially observed. Transmission rates estimated using these partially observed networks would be overestimated compared to reality due to the lower number of contacts, which could lead to an incorrect evaluation of the impact of interventions [25–27]. By comparison, although they do not provide individual-level information, contact matrices and summary statistics such as contact rates between individual groups are more readily available, as they can be inferred using simple cross-sectional survey data [28–30].

Here, we propose an algorithm to stochastically reconstruct realistic contact networks from partially observed contact data in healthcare settings (HCS). To validate our approach, we use close-proximity data collected in a long-term care facility (LTCF) during the i-Bird study [17,31]. We first illustrate the typical complexity of contact structures in HCS through the i-Bird network example. We then compute summary contact parameters from these data to generate reconstructed contact networks and compare these synthetic contact networks with the observed data.

## Methods

### Building synthetic contacts in a HCS

#### Algorithm outline

We built an algorithm to stochastically reconstruct a realistic full temporal network of inter-individual close-proximity interactions (CPIs, at less than 1.5m) in a HCS using parameters estimated from empirical individual contact data. This algorithm generates a new synthetic network which notably reconstructs contacts at times when individuals were known to be present in the HCS but had no contact data recorded, which we consider to be a recording bias. The synthetic network hence includes both the observed and unobserved parts of the empiric network. This approach first involves the calculation of contact rates and durations between individuals, stratified by the individuals’ ward, category (patient, or staff profession), type of day (weekday or weekend) and hour. The algorithm then reconstructs a new network, taking as input these summary statistics as well as data on presence days for each individual in the facility. Each CPI is generated stochastically, with individuals chosen in order to promote recurring contacts, based on a probability estimated from the data.

#### Estimation of contact rates from the data

Contact rates per hour (*h* from 00h to 23h), category of individual (*C_i_*, i.e. patient, or hospital staff profession) and ward *W_i_* are estimated from the data as:

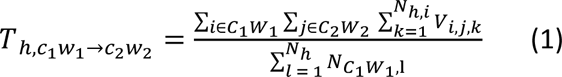

where *T_h,c1w1→c2w2_* is the average per-person contact rate at the hour *h* between individuals from category *C_1_* belonging to ward *W_1_* and individuals from category *C_2_* belonging to ward *W_2_*. For given hour *h* and individual *i*, *N_h,i_* is the number of instances of the hour *h* where at least one contact was recorded for individual *i*. For example, if *i* had a contact recorded on Tuesday 11^th^ August at 10h, and on Tuesday 18^th^ August at 10h, *N_10,i_* would be equal to 2. For two individuals *i* from C1W1 and *j* from C2W2, *V_i,j,k_* indicates whether contacts have been recorded between them on instance *k* of the hour *h*: it equals 1 if *i* and *j* had at least one contact recorded at that time, and 0 otherwise. Finally, *N_h_* is the total number of instances of the hour *h* in the full dataset and, for a given instance *l* of the hour *h, N_C1W1,l_* is the number of individuals from *C_1_W_1_* that had any contact recorded during that hour.

This estimation is conducted separately for contacts during weekdays and contacts during weekends.

#### Estimation of recurring contacts

For each individual *i*, we calculate the probability of recurring contact for each day *d* between the first (*d_0_*) and last (*d_max_*) days where a contact was recorded for *i*, according to

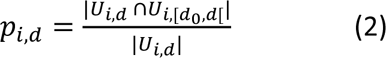

Where *U_i,d_* is the set of unique individuals with whom *i* had a contact on day *d*, *U_i,[d0,d[_* is the set of unique individuals with whom *i* had at least one contact on any day between the first day *d_0_* and the current day *d* (*d* non-included), and the notation |*x*| indicates the cardinal of the set *x*. For example, if *i* had a contact with four unique individuals on day *d*, and previously had a contact with two of those on any day between *d_0_* and *d*, the probability of recurring contact for day *p_i,d_* would be 2/4 = 0.5.

We then calculated the mean daily probability of recurring contacts for individual *i* across all days as

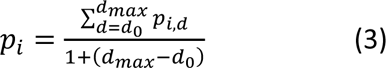

Finally, we calculated the mean probability of recurring contacts by individual category *c* (patient or staff) as

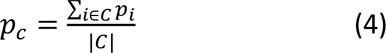

Where *C* represents the set of individuals belonging to category *c*.

#### Generation of synthetic CPIs: number and identity of individuals in contacts

For each hour of our period of interest, we estimate the number of contacts between individuals present in the HCS during that hour, determined using admission data and staff schedule. We generate the number of individuals *n* from category *C_2_S_2_* in contact with an individual *i* from category *C_1_S_1_* during an hour *h* by sampling from a Poisson distribution with the mean being the contact rate as described above. Before selecting these *n* individuals, since contacts are generated dynamically, we check if *i* is already included in the contacts of individuals from *C_2_S_2_* during *h*. If *n’* individuals from *C_2_S_2_* have already had a contact with *i* during *h*, we only select *n-n’* new individuals from those available, in order to avoid double counting.

These individuals are selected by favouring contacts between individuals who have already met at any other time previous to *h*. Let *p_c_* be the probability of a recurring contact for category *c* (patient or staff) of the individual *i*. To determine the identity of the *n* individuals in contact with *i*, we draw a random number *r* ∼ *Uniform(0,1)*

- If *r* ≤ *p_c_*, a recurring contact is generated: *j* is chosen among *S*, the subset of *C_2_S_2_*
individuals who previously met *i*, according to probability *p_i→j_*:

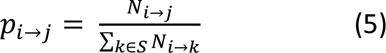

Where *N*_*i*→*j*_ is the number of previous contacts between *i* and *j* before hour *h*, and ∑_*k*∈*S*_ *N*_*i*→*k*_ is the number of previous contacts between *i* and each individual *k* belonging to *S*.
- Otherwise, the contact is not recurring: the individual *j* in contact is randomly and uniformly chosen among *S’*, the subset of *C_2_S_2_* individuals who have not yet met *i*.

#### Generation of contact durations

For each contact between two given individuals *i* from *C_1_S_1_* and *j* from *C_2_S_2_* the duration of contact is sampled from a log-normal distribution calibrated from the observed mean and variance of contact durations between individuals from *C_1_S_1_* and *C_2_S_2_* on hour *h*.

### Validation dataset: the i-Bird network

#### Dataset description

We validate our algorithm by applying it to data collected during the Individual-Based Investigation of Resistance Dissemination (i-Bird) study [17,31]. This study took place in a rehabilitation and long-term care facility (LTCF) from the beginning of July to the end of October 2009. Over this period, each participant (patient or hospital staff) was wearing an RFID sensor that recorded CPIs every 30 seconds. Here, we only used contacts recorded between 27 July to 23 August 2009 (included). This period corresponds to the weeks between two sensor battery replacements and hence avoids interference due to loss of contact. A temporal network of proximities was therefore available over 28 days with information on individual ID and ward of affectation.

The LTCF was structured into five wards: three neurological wards, one nutritional care ward and one geriatric ward. Patients were systematically linked to a ward, whilst some staff were mobile and not linked to a specific ward. For the purpose of this work, we considered here that mobile staff belonged to an “artificial” 6^th^ ward, to compute contact rates according to the algorithm detailed above. Staff were divided into 13 professions: administrative, animation/hairdresser, logistic, hospital service agent, porter, occupational therapist, physiotherapist, other rehabilitation staff, nurse, head nurse, care assistant, medical student/resident, and physician. A total of 200 patients and 213 hospital staff were included and had contacts recorded during the 28 days of study.

We used hospital staff schedules to determine the hourly presence of each staff and compared these schedules to the dates and times when staff had any contact recorded. We assumed that, in reality, staff would have at least one contact with any other individual during any given hour of their presence time, hence if no contact was recorded for a given hour of presence we considered this was missing data rather than true absence of contact. Through this, we estimated that the median percentage of a staff’s total presence time when no contact data was recorded was 40.0% (interquartile range (IQR): 0-75.0%). We repeated this analysis for patients at the daily instead of hourly level, as we only had access to admission and discharge dates for patients. We estimated that the median time when no contact data was recorded was 33.3% (interquartile range (IQR): 10.5-53.6%) of a patient’s presence days.

Although the overall compliance was high (90% of individuals agreed to wear a sensor), there was therefore substantial heterogeneity in the individual coverage of the raw i-Bird network (Supplementary Figure 1). Interestingly, there was no correlation between the proportion of presence time during which contact data were recorded for a given individual and their average number of contacts on presence days where data were available, nor their total presence time (Supplementary Figure 2).

#### Observation bias process

As mentioned earlier, the observed i-Bird network, as any real-life data, includes recording biases leading to some periods of non-recording of CPIs, with the extent of this bias varying between individuals. To make our reconstructed networks comparable to the observed one, we therefore introduced an observation bias process. For each individual in the observed network, we identified the hours with no contact recorded. We then removed those individuals on those hours before proceeding with the algorithm described above. The resulting “reconstructed biased network” and the observed network hence suffer from the same bias and are comparable.

### Simulations and analysis

From the analysis of the i-Bird empiric network and data, we used our algorithm to generate 100 full synthetic reconstructed networks, and 100 reconstructed networks with observation bias. For comparison, we also generated 100 pseudo-random contact networks with observation bias, and 100 without. The latter networks simulate contacts without taking into account the ward, staff category, and probability of recurring contact in the calculation of contact rates and durations. The patient-patient, staff-staff, and patient-staff contact rates are calculated as detailed in the section “Estimation of contact rates from the data”, treating all staff as if they were part of the same profession, and all individuals as if they were part of the same ward. At each contact, the individual encountered is therefore chosen randomly from all those present in the LTCF at that time, regardless of whether or not the individual was previously encountered.

We implemented the algorithm in C++ with the repast HPC 2.3.0 library. All simulations were performed on the Maestro cluster hosted by the Institut Pasteur. The networks were analysed in R [32], using the igraph package [33]. The relevant contact networks and analysis code are available in the following GitHub repository: https://github.com/qleclerc/network_algorithm.

### Validation of the full reconstructed networks

For validation, we also applied the algorithm to each of the 100 reconstructed networks with bias, considering them as empiric networks. This allowed us to generate 100 new full reconstructed networks from fully known networks, and confirm these “re-simulated networks” were similar to the full reconstructed networks generated from the observed data.

## Results

### Description of HCS contact heterogeneity: the example of the i-Bird dataset

In this section, we illustrate the typical complexity of contact structures in HCS using the i-Bird network. While the algorithm makes use of data at the hourly level, in this section the contact data are aggregated at the daily level, so that if two individuals have two separate contacts with each other at different times of the day, this is only counted once. The contact network is considered undirected, since contacts are assumed to be reciprocal. Daily-averaged contact matrices built from these data are described in a previous work [31].

We first summarise the observed temporal network recorded in the LTCF during the i-Bird study, comparing the total daily network and subgraphs with only patient-patient, staff-staff, or patient-staff contacts (Figure 1a-d). Table 1 provides the degree, global efficiency, density, transitivity, assortativity and temporal correlation of these four networks. The mean degree of the total network per day is 12.99 (standard deviation: 3.53), which corresponds to the average number of unique contacts per individual per day. In the subgraphs, the degree is highest in the patient-staff subgraph (8.09; sd: 1.89), although we still note a relatively important number of patient-patient contacts, with a degree of 5.25 (sd: 1.87) in the corresponding subgraph. The distribution of individual degrees for all individuals and all days across the total network is heterogeneous, with a squared coefficient of variation equal to 0.44 (Figure 1e). The global efficiency of the total network is 0.40 (sd: 0.05), meaning that on average the shortest path between any two individuals has a distance of 2.5 (whereby the shortest path between two individuals in direct contact would be of distance 1). As expected, the efficiencies are lower in the subgraphs, since we remove individuals and hence increase the distance between those remaining (patient-patient: 0.25 (sd: 0.08, distance: 4); staff-staff: 0.32 (sd: 0.10, distance: 3.1); patient-staff: 0.31 (sd: 0.05, distance: 3.2)). Densities in the total network and subgraphs are relatively low (< 0.1), indicating that less than 10% of all possible connections between individuals in the network are actual observed connections.

**Figure 1:**
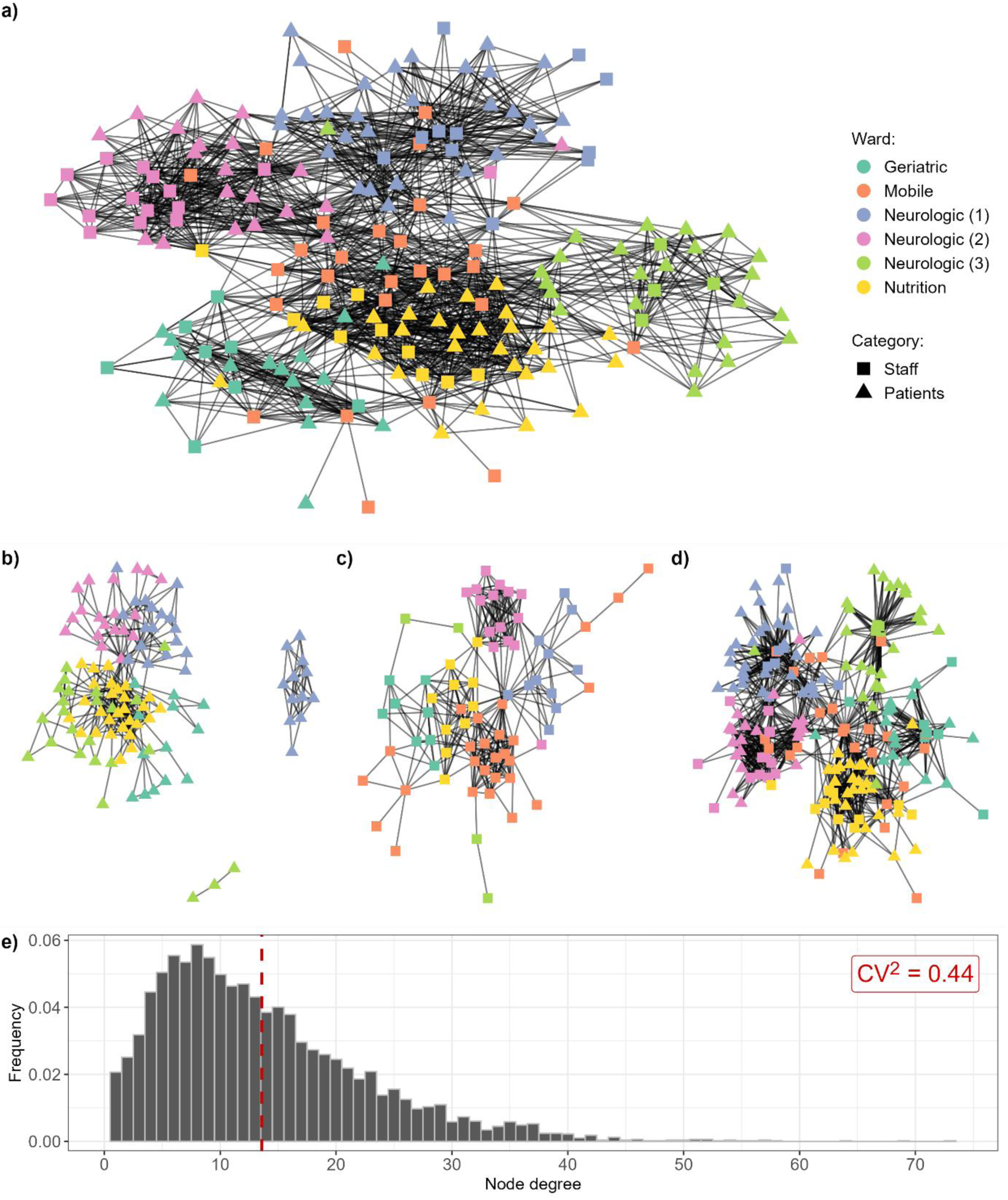
Representation of the observed network recorded during the i-Bird study: **(a) total network, and (b) patient-patient, (c) staff-staff and (d) patient-staff subgraphs on a single day.** The date of 28^th^ of July 2009 was chosen arbitrarily. The layout was calculated using the Kamada-Kawai algorithm, with no weights applied to edges. **e) Distribution of individual degrees for the total network per person per day, across the entire study period.** The dashed red line indicates the mean degree (13.59). CV: coefficient of variation (standard deviation/mean).

**Table 1:** Summary of network characteristics for the observed i-Bird total network, patient-patient subgraph, staff-staff subgraph, and patient-staff subgraph. Values were estimated for each day of the 28-days period and summarised here with the mean and standard deviation (sd). Transitivity is not calculated for the patient-staff subgraph as triangles of contacts cannot occur in this network.

|  | Total | Patient-patient | Staff-staff | Patient-staff |
| --- | --- | --- | --- | --- |
| **Degree (sd)** | 12.99 (3.53) | 5.25 (1.87) | 5.82 (1.87) | 8.09 (1.89) |
| **Global efficiency (sd)** | 0.40 (0.05) | 0.25 (0.08) | 0.32 (0.10) | 0.31 (0.05) |
| **Density (sd)** | 0.07 (0.01) | 0.05 (0.01) | 0.09 (0.01) | 0.05 (0.00) |
| **Transitivity (sd)** | 0.37 (0.02) | 0.41 (0.05) | 0.56 (0.07) | NA |
| **Assortativity (sd)** |  |  |  |  |
| ***By degree*** | -0.13 (0.10) | 0.22 (0.10) | 0.14 (0.14) | -0.42 (0.14) |
| ***By ward*** | 0.59 (0.08) | 0.77 (0.11) | 0.72 (0.09) | 0.47 (0.09) |
| **Temporal correlation** | 0.47 (0.11) | 0.65 (0.07) | 0.35 (0.16) | 0.41 (0.12) |

Transitivity in the total network is high (0.37; sd: 0.02), meaning that for any two individuals *a* and *b* both in contact with the same third individual *c*, the probability that *a* and *b* are also in contact is 0.37. Transitivity is also high in the patient-patient and staff-staff subgraphs, but this metric is not relevant for the patient-staff subgraph – it is impossible for a triangle of contacts to occur in this subgraph as it excludes staff-staff and patient-patient contacts by design. Assortativity by degree is negative in the total network (-0.13; sd: 0.10), indicating that highly connected individuals are more likely to be in contact with less connected individuals. It is also strongly negative in the patient-staff subgraph (-0.42; sd: 0.14), reflecting the expected disassortivity of healthcare contacts, where each staff member is in contact with multiple patients, whilst each patient is contact with relatively few staff members. In the patient-patient and staff-staff subgraphs, assortativity by degree is positive, as frequently seen in social networks.

Visually, we observe that contacts are naturally clustered by ward (Figure 1a-d). This is reflected in the assortativity by ward, which is systematically high (> 0.45) and indicates that individuals in a ward are always more likely to have contacts with other individuals in the same ward than with individuals in other wards (Table 1). We also observe that contacts exist between all grouped staff professions and patients in different wards, although the distribution is heterogeneous (Figure 2a-b). For example, the median number of wards with which a care assistant (orange) is in contact with is two, while almost all porters (yellow) have contacts with patients from all five wards (Figure 2b).

**Figure 2:**
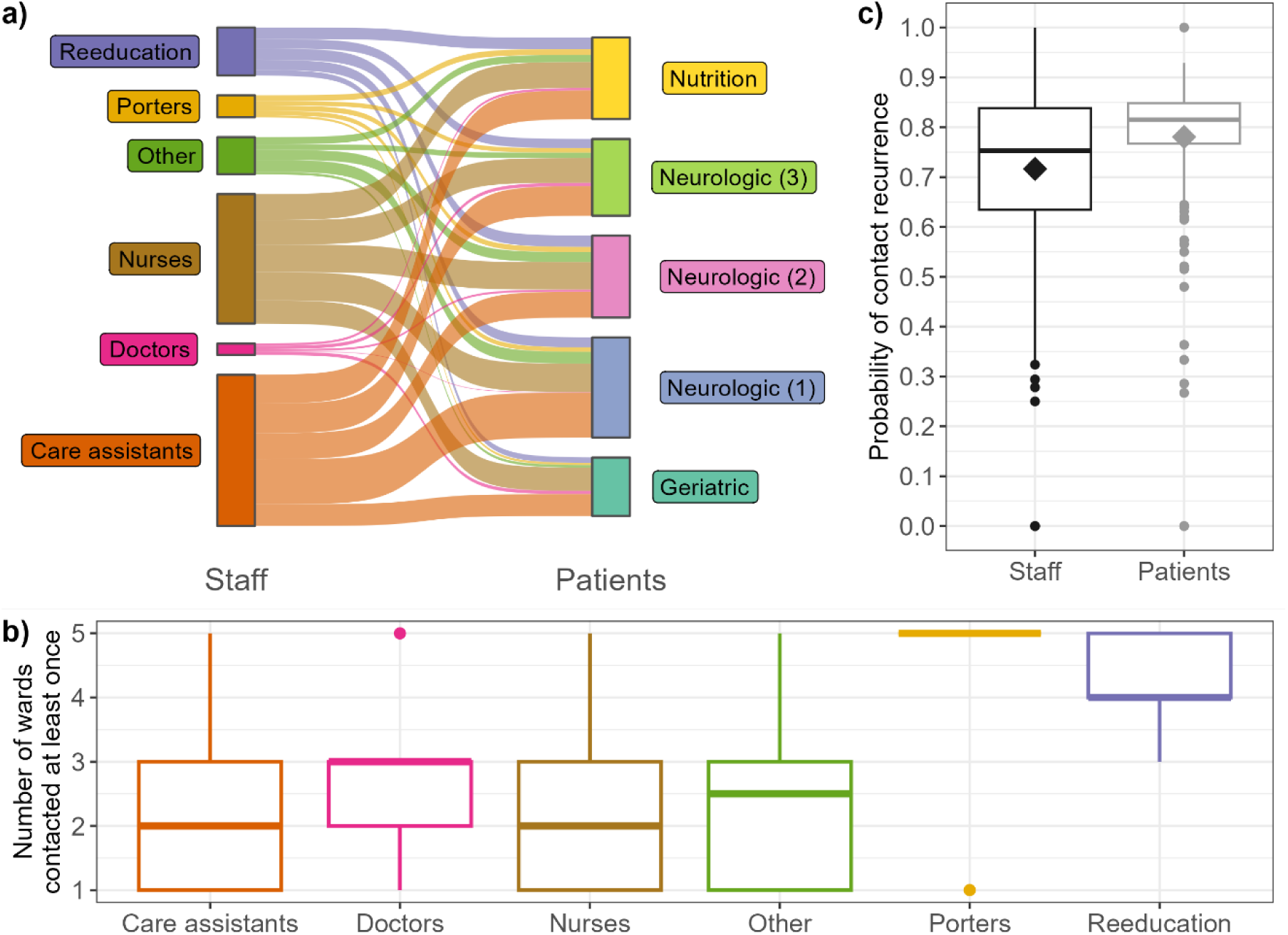
Description of contact heterogeneity and recurrence across the facility. **a) Repartition of contacts between grouped staff professions and patient wards.** A link between one staff category and one patient ward indicates that, at any point during the investigation period, a staff member from that category had a contact with a patient from that ward. For ease of visualisation, occupational therapists, physiotherapists, and other re-education staff are grouped into “Reeducation”; administrative, animation/hairdresser, logistic, and hospital service agents are grouped into “Other”; and nurses, head nurses, and students/interns are grouped into “Nurses”. Porters, doctors and care assistants are not grouped. **b) Distribution of number of wards with which each staff member has had at least one contact with during the study period. c) Distribution of probabilities of recurring contacts.** Each observation is calculated over the entire studied period, and corresponds to the average probability for one staff or one patient to form a new contact with a previously-met individual (staff or patient) over the studied period rather than a new individual. Diamonds indicate the mean values.

Overall, contacts are relatively well maintained over time, as shown by the temporal correlation coefficient of 0.47 (sd: 0.11, Table 1). This corresponds to the average probability that, between two subsequent days, an individual maintains the same number of unique contacts, with the same individuals. This metric is highest in the patient-patient subgraph (0.65, sd: 0.07) and lowest in the patient-staff subgraph (0.35, sd: 0.16), indicating that patients tend to have the same contacts with each other every day, whilst contacts amongst healthcare workers often vary between subsequent days. This consistency over time is reflected in the high probability of recurring contacts (mean probability: 0.78 for patients, 0.71 for staff), although we note more variability amongst staff than patients (Figure 2c).

All the characteristics described above differ between weekdays and weekends in the network and indicate that there are fewer contacts during weekends (Supplementary Table 1). This difference is reflected in the temporal correlation, which tends to be high when comparing Sunday to Saturday, but low when comparing Saturday to Friday and Monday to Sunday, indicating that the structure of the network changes the most between these timepoints (Supplementary Figure 3).

### Comparison of synthetic and observed networks

To illustrate and validate our algorithm, we applied it to the i-Bird network described above to stochastically construct four types of synthetic networks using the estimated contact parameters: 100 full reconstructed networks, 100 reconstructed networks incorporating observation bias, 100 full pseudo-random networks, and 100 pseudo-random networks incorporating observation bias. We expected that the characteristics of the reconstructed networks with observation bias would be broadly similar to those of the observed i-Bird network. Summary network characteristics are reported in Figure 3 and Supplementary Figure 4.

**Figure 3:**
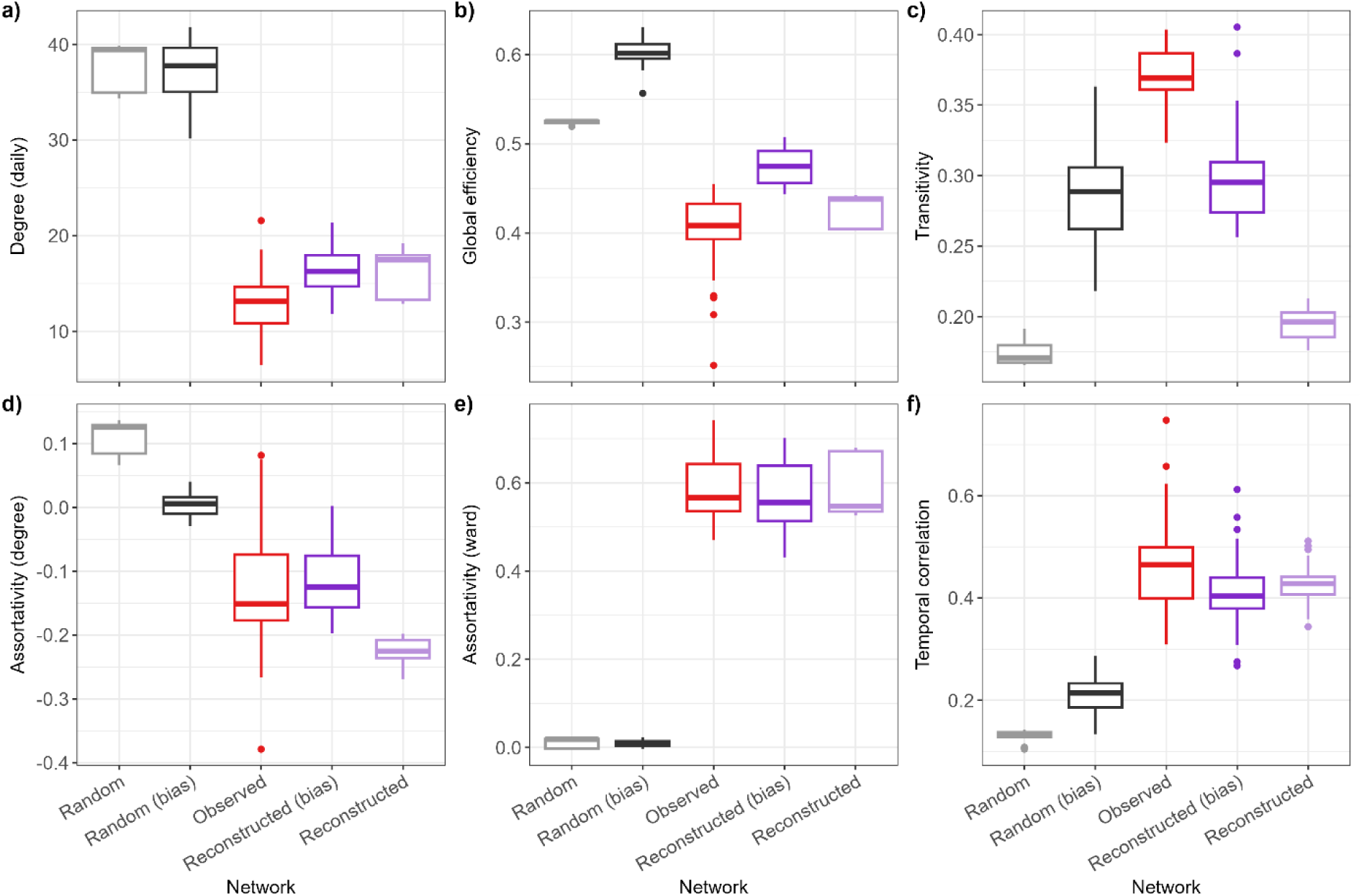
Comparison of network characteristics. The reconstructed networks with observation bias exclude individuals from the network at times when they were known to not wear their sensors. The random networks did not take into account the ward-level structure of the contacts or the probability of recurring contacts. Boxplots for the observed network show the distribution of values calculated for each day. Boxplots for all reconstructed and random networks show the distribution of the median values calculated for each day across 100 networks.

The daily degrees in the reconstructed networks were slightly higher than the observed network (Figure 3a). Global efficiency was similar between the observed and reconstructed networks, but slightly higher in the reconstructed network with bias (Figure 3b). This is because the algorithm with bias removed individuals from the network at times when they did not wear their sensor during the study, hence reducing the average distance between remaining individuals. For the same reason, the density of the reconstructed network with bias was slightly higher than the observed (Supplementary Figure 4). Transitivity was slightly higher for the reconstructed network with observation bias than without, but lower than the observed network in any case (Figure 3c), as expected since the algorithm did not take into account any element of transitivity when constructing synthetic networks. Finally, assortativity by degree and by ward, as well as temporal correlation, were all well preserved in the reconstructed networks (Figure 3d-f). As a comparison, the random networks with or without bias either substantially over- or under-estimated the values for all metrics compared to the observed network (Figure 3a-f), although we note that transitivity was similar to the other synthetic networks (Figure 3c).

The hourly distributions of numbers of unique patient-patient, staff-patient and staff-staff contacts in the reconstructed network with bias align with those in the observed network (Figure 4a). Whilst these two networks are only partially observed since individuals in the i-Bird study did not have contacts recorded during all their presence days, those unobserved contacts are present in the reconstructed network without bias, leading to approximately twice as many contacts in that network (Figure 4a). Similarly, the random network without bias which is only informed by the hourly distribution of patient-patient, staff-staff and patient-staff contact rates is aligned with the reconstructed network (Figure 4a).

**Figure 4:**
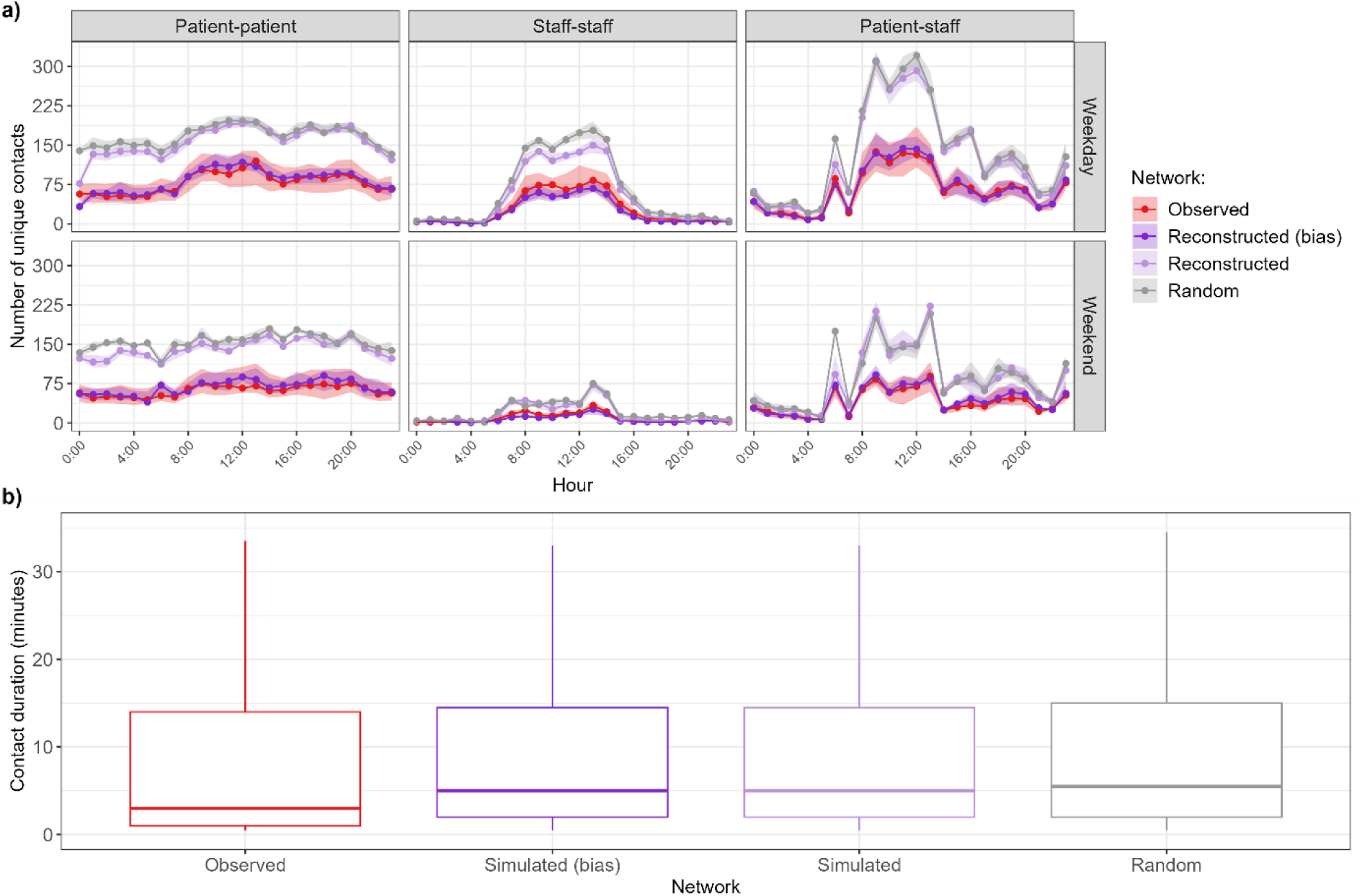
Comparison of network contact number and duration. **a) Distribution of number of unique contacts per hour, separated by type of day (weekday or weekend).** Points correspond to the median, and the shaded areas correspond to the interquartile range. **b) Distribution of contact durations.** For ease of visualisation, outliers are not shown on the graph.

Finally, the distributions of contact durations in the synthetic networks were similar to the distribution in the observed network, although there were slightly less contacts with short durations (Figure 4b). This is because all networks sample their contact durations from a lognormal distribution parameterised by the mean and variance estimated from the data, which puts less emphasis on very short contacts of less than one minute (Supplementary Figure 5).

In supplementary analyses, we assessed the robustness of our algorithm by quantifying the variability of network characteristics across 100 reconstructed networks without bias (Supplementary Figure 6). The variability across reconstructed networks was not statistically significant for any metric (Kruskal-Wallis test, p value > 0.05) except for assortativity by degree (p < 0.001). We also aimed to validate our approach by generating “re-simulated” networks informed by summary statistics derived from the reconstructed networks with bias. These re-simulated networks are similar to the full reconstructed networks, indicating that our algorithm consistently recreates realistic networks and reconstructs unobserved contacts (Supplementary Figure 7). However, the number of patient-patient contacts in the re-simulated networks is slightly higher than in the reconstructed networks (Supplementary Figure 7).

## Discussion

### Summary of findings

In this article, we present an approach to construct stochastic synthetic temporal contact networks in HCS from partially observed contact data. The i-Bird network illustrates the typical complex contact structures in HCS, notably with a strong assortativity by ward, varying contact rates between different staff categories and patients, and different contact structures on weekends compared to weekdays. Importantly, we observed temporal correlation between subsequent days in the network, and we estimated that individuals were generally more likely to have contacts with other individuals they previously met rather than new individuals. Our reconstruction algorithm successfully captured the heterogeneity of the observed network by taking into account contact rates by hour, type of day (weekday or weekend) and staff category, and probabilities of recurring contacts estimated for patients and staff. The resulting reconstructed networks reproduced well the characteristics of the observed network, as well as the specific distribution of unique contacts per hour.

The value of approaches to stochastically generate realistic contact networks has been previously discussed for schools or workplaces [6,34], although the complexity of the contact structures in those settings is arguably lower than what we observed here. While these approaches extended networks by either repeating contact structures at fixed intervals or randomly shuffling links [6,34], our algorithm dynamically and stochastically constructs new contacts at each hour based on the empirical contact rates. Previous algorithms also attempted to reconstruct missing contacts for non-participants [35]. While this was not accounted for here (e.g. visitors, see below for details), here we conduct this reconstruction at a higher resolution, since in reality participating individuals can also have contact data missing only for some hours or days of their total presence time. Finally, the novelty of our approach here is that we conduct a direct comparison between the output of our algorithm and the observed contact network, as opposed to other algorithms which attempted to build networks directly from contact diaries and hence did not have access to an observed network for comparison [36].

### Similarities between the observed and reconstructed networks

Although 90% of individuals agreed to wear a sensor during the study, the i-Bird contact network was only partially observed, since the median time when contacts were not recorded was 33.3% (IQR: 10.5-53.6%) of a patient’s presence days (40.0%, IQR: 0-75.0% for staff). This could have occurred for a number of reasons which we cannot distinguish, including depleted batteries, sensor malfunction, imperfect sensor-wearing compliance, or temporary patient releases from the facility (see Limitations below). However, since the average contact rates of individuals did not correlate with the proportion of their presence time during which no contact data were recorded (Supplementary Figure 2), it can be assumed that contact patterns during unobserved times were similar to those on observed times. With that assumption, we were able to reconstruct contacts at those times when individuals were present but had no reported contact data. The resulting full reconstructed network is a valuable representation of individual interactions, as it represents the “true” contact network, compared to the i-Bird empirical network which was only partially observed. Although we were inherently limited in our ability to validate this network since the real, fully observed network was not available, we compared it to a re-simulated network which used the reconstructed network with observation bias as input. The reconstructed and re-simulated networks without bias were almost identical with regards to all the network metrics we considered (Supplementary Figure 7), demonstrating the consistency of our algorithm to reconstruct contacts.

The reconstructed network with bias and the observed network had similar positive assortativity by ward, as expected since the input data captured the contact structure by ward. The negative assortativity by degree was also similar, however we noted variability between different networks generated independently by the algorithm (Supplementary Figure 6). Since the algorithm did not directly account for assortativity when simulating networks, this similarity stems from our use of a recurring contact probability coupled with the contact rates estimated by staff categories, resulting in a non-random contact structure with regards to this metric. The hourly contact distribution of patient-patient, staff-staff, and patient-staff contacts was also successfully reproduced by our algorithm.

A key metric of interest here is temporal correlation, which indicates how conserved the network structure is over time. This type of metric is useful to determine the efficiency of disease spread across temporal networks over time [37–40]. Since our algorithm took into consideration the probability of recurring contacts between individuals, our reconstructed networks displayed similar temporal correlation as observed, whilst random networks substantially underestimated this. This aspect is therefore an important strength of our approach, compared to only using estimated average contact rates to construct synthetic contacts.

### Limitations of the algorithm

Density and global efficiency in the reconstructed network with bias were slightly higher than in the observed network. This is a likely consequence of our observation process which forcibly removed individuals from the network at times when they had no contacts recorded, hence reducing the number of nodes available in the network. Simultaneously, there was still a need at those times to generate some novel contacts between individuals who never previously met, since the probability of recurring contacts was less than 1. Combined, these elements increased the overall connectivity amongst all individuals in the reconstructed network with bias. Although this could facilitate disease transmission across these reconstructed networks if they are used for such purpose [41], the high assortativity by ward may counter this effect by slowing down transmission across the entire healthcare facility.

Our algorithm did not specifically account for transitivity when recreating contacts. This is likely why the resulting transitivity was similar to that of the random network and underestimated the observed value (Figure 3). Similarly to density and global efficiency mentioned above, any transitivity in the reconstructed network was likely an indirect consequence of assortativity by ward, restricting the pool of available individuals to generate contacts and leading to interconnectivity between individuals present in the same ward. Whilst we could extend our algorithm to consider transitivity when choosing the individuals to put in contact, we decided not to do this here to maximise the generalisability of our approach by not requiring such highly detailed contact data. In any case, this may not substantially affect disease transmission simulated across these networks, since previous work has shown that transitivity is a poor predictor of the total number of individuals who would be infected across the network [41].

Although our algorithm can capture individual presence and absence times, information about patient temporary releases from the LTCF (e.g., for weekends with their families, or for shopping outside) was not available in the i-Bird data, hence such events were not accounted for here, although they may occur frequently in a LTCF. Consequently, the number of presence days/hours may have been overestimated, leading to an overestimation of contact days among patients. Although this is negligible when comparing the observed and reconstructed network with bias, this is likely why the re-simulated networks slightly overestimated the number of patient-patient contacts compared to the full reconstructed network (Supplementary Figure 7). We expect that this overestimation would be absent in HCS with more complete information on individual presence, or in acute care facilities with shorter patient lengths of stay and where temporary releases are less common. Similarly, our algorithm does not consider the contacts of visitors in the hospital, and we did not have data in the i-Bird study on visitors which we would have required to validate the synthetic networks. Consequently, our description of the contact structure in the LTCF is not exhaustive, although this does not affect the ability of our algorithm to reproduce patient-staff contacts.

When reconstructing missing contacts, we assumed that if a staff member (patient) was present in the facility at a given time but did not have any contact with anyone else recorded at that hour (day), this represented unobserved data. In reality, there may be rare instances where individuals truly did not have any contact with anyone else over a time period. In such instances, our algorithm would over-estimate contacts by forcibly reconstructing contacts for those individuals at those times. However, we expect this would only occur at times with limited contact rates (e.g. during the night), therefore the empirical contact rates would be small and only a couple of contacts may be erroneously reconstructed by the algorithm.

### Future work

In this study, we show that our algorithm can accurately reproduce the contact structure using as input contact data from a given long-term care facility. A first important next step would be to repeat this analysis using data collected in a different HCS such as acute care, over a different time period. This is because contact structures are known to vary between different HCS such as long-term or acute, with more/less contacts between different individual categories, varying recurring contact probabilities etc. Similarly, even though we tested our algorithm using substantial data covering four weeks, this contact structure may not be representative of other time periods. Notably, the i-Bird data we used was collected in the middle of the summer, which is a holiday period in France and may have affected contact patterns. Although we do not expect that our algorithm will perform differently since it has been designed to be generalisable, the strengths and limitations we have highlighted above may be more or less relevant in these different settings. For example, in a setting with low transitivity, the fact that our algorithm underestimates this metric would be less problematic.

Here we directly re-used patient admission and discharge data as well as staff schedules to identify which individuals were present in the facility at each hour, and hence whom the algorithm had to build contacts for. While this choice was coherent since our aim was to compare the observed and reconstructed networks, a second possible extension of our work would be to simulate the presence of individuals over time. This could be implemented by extracting admission and discharge rates for each category of staff and patients and using these values to recreate new presence times for individuals by sampling from relevant probability distributions while maintaining constrains on each population size. This would allow us to further account for possible variability in the structure of the population in the facility, add flexibility in building synthetic networks for settings where this data may not be fully available, and hence add further stochasticity in our algorithm.

Since contact data may only be available for short periods of time (e.g. a few days [22]), a third question of interest would be to understand the volume of data required to generate realistic temporal contact networks using our algorithm. In our main analysis, we used the entire four weeks available to both derive contact parameters and compare the reconstructed and observed networks. For sensitivity, we also considered smaller timer periods to calculate the summary contact parameters required by the algorithm (Supplementary Figure 8). As expected, this led to variability amongst the reconstructed networks depending on the length of the period used, since this reduced the number of data points used to estimate the average contact rates used by the algorithm. In any case, the main risk of using only a short period of time is to miss out on some contacts between categories. For example, during a single week, by chance there may not be any observed contact between patients from one ward *w1* and a nurse from another ward *w2*, while in reality over a longer period of time we may observe a few of such contacts. In that case, the algorithm will systematically assume that such contacts never occur during the entire period over which the reconstructed networks are generated and will therefore construct an incomplete network. A further extension of our algorithm could include the possibility of creating such unobserved links, but this would still require either assumptions or information on the nature of those links. Therefore, it is essential for users to be confident that the data they use include contact rates for all relevant categories in their setting and for typical representative days.

As discussed above, taking into account the probability for contacts to be recurring instead of assuming a uniform distribution is a key element of our approach. Here, we estimate the average probabilities of recurring contacts in the studied LTCF over the studied period as 0.71 for staff and 0.78 for patients, but we note some individual variation in this value (Figure 2, interquartile range for staff: 0.63-0.84, for patients: 0.77-0.85). In addition, our estimation here is made using the entire observed contact networks over the study period, but this may be difficult in instances where only limited data are available. For sensitivity, we investigated the impact of manually setting the probabilities to 0.1, 0.5 and 0.9 for both staff and patients (Supplementary Figure 9). This led to important variations in assortativity by degree and temporal correlation compared to using the estimated probability. A greater understanding of this recurring contact probability in various settings would be key to better understand contact formation and heterogeneity, and could be directly taken into consideration in our algorithm since it has been designed to use this probability. In healthcare settings, this probability could likely be estimated without requiring complete contact data, using information on staff schedules and patient ward or room allocation instead.

Finally, other methodological approaches could be considered to reconstruct realistic contact networks. For example, deep learning algorithms such as graph convolutional networks (GCN) have become increasingly popular for this purpose, particularly in the context of infectious disease transmission [42–46]. It would be interesting to compare the performance of these approaches with our algorithm to estimate network characteristics and reconstruct unobserved contacts. However, traditional GCN approaches do not account for temporal dependencies between contacts such as the ones we observed in the i-Bird network where the probability of recurring contacts plays a key role [47,48]. On the other hand, temporal graph networks can capture this temporal dependency [49,50], but require substantial computational resources to be applied to a network such as i-Bird, with hundreds of interactions recorded every 30 seconds during several weeks. Finally, deep learning methods require large amounts of training data. Democratising their use would therefore first require new studies to collect close-proximity interaction data in different settings and time periods, presenting further logistical challenges.

### Implications

Our algorithm relies on computing summary statistics from an observed network, then using these statistics to stochastically reconstruct contact networks. Such statistics can be derived directly from other observed networks, as we have done here to validate our approach. In that case, instead of only relying on a single observed network, our approach provides multiple realistic reconstructed networks enabling to consider the impact of stochasticity of the contact structure and on subsequent epidemic risk in a given setting. Our approach, by providing data augmentation, also enables to infer information on potentially unobserved contacts and generate extended realistic temporal dynamics over longer time periods than the period of data collection.

Alternatively, summary contact statistics could be more simply collected from cross-sectional surveys or even derived exclusively from individual schedules, which would not require a detailed and costly follow-up using sensors. In this scenario, the only other data required would be individual presence times, which should either be routinely available (e.g. in healthcare settings or schools) or relatively easy to collect (e.g. in workplaces). Although as mentioned in the Limitations, the amount of data our algorithm requires to generate realistic networks is still unclear, our approach could ultimately be used to generate contact networks from contact matrices. This would substantially facilitate research on the impact of contact heterogeneity in various populations and settings, as others have previously discussed [36].

In conclusion, our algorithm can generate temporal contact networks in a healthcare setting by taking into consideration empirically measured contact rates based on close-proximity sensors, as opposed to most available packages which only construct static networks and rely on hyperparameters [51,52]. These temporal networks can then be analysed with mathematical models to evaluate the potential impact of interventions against disease transmission [11–14]. In particular, this will improve the wider applicability of individual-based model which make it possible to account for detailed contact heterogeneity in testing the effect of interventions targeting highly specific individuals.

## Data Availability

The relevant contact networks and analysis code are available in the following GitHub repository: https://github.com/qleclerc/network_algorithm

## Acknowledgments

AD, LT and LO received funding from the French National Research Agency (SPHINX-17-CE36-0008-01). DG received funding from the National Clinical Research Program and the Investissement d’Avenir program, Laboratoire d’Excellence "Integrative Biology of Emerging Infectious Diseases" (ANR-10-LABX-62-IBEID). The authors would like to thank Eric Fleury, Pierre-Yves Boëlle, Vittoria Colizza and Pascal Crépey for helpful discussions on the analysis of the contact network.

