## Supplementary Material for "An algorithm to build synthetic temporal contact networks based on close-proximity interactions data"

**Short title: An algorithm to build synthetic contact networks from contact data**

**Audrey Duval[1,2,3+¤], Quentin Leclerc[1,2,3+], Didier Guillemot[1,2,4], Laura Temime[3,5#], Lulla Opatowski[1,2#]**

[+] these authors contributed equally

[#] these authors contributed equally

[1]Institut Pasteur, Université Paris Cité, Epidemiology and Modelling of Bacterial Escape to Antimicrobials (EMEA), Paris, France

[2]INSERM, Université Paris-Saclay, Université de Versailles St-Quentin-en-Yvelines, Team Echappement aux Anti-infectieux et Pharmacoépidémiologie U1018, CESP, France

[3]Laboratoire Modélisation, Epidémiologie et Surveillance des Risques Sanitaires (MESuRS), Conservatoire National des Arts et Métiers, Paris, France

[4]AP-HP, Paris Saclay, Department of Public Health, Medical Information, Clinical research, F-92380, Garches

[4]Institut Pasteur, Conservatoire National des Arts et Métiers, Unité PACRI, Paris, France

¤ Current address : Imagine Institute, Data Science Platform, INSERM UMR 1163, Université de Paris, Paris, France

**Keywords:** long-term care facility, contact network, close-proximity interactions, sensors, network reconstruction

### Supplementary Material

**Supplementary Table 1: Summary of network characteristics for the observed total network, patient-patient subgraph, staff-staff subgraph, and patient-staff subgraph, separated by weekday or weekend.** Values were estimated per day, with mean and standard deviation (sd) presented here. Transitivity is not shown for the patient-staff subgraph as triangles of contacts cannot occur in this network.

| | **Total** | | **Patient-patient** | | **Staff-staff** | | **Patient-staff** | |
|---|---|---|---|---|---|---|---|---|
| **Day type** | **Weekday** | **Weekend** | **Weekday** | **Weekend** | **Weekday** | **Weekend** | **Weekday** | **Weekend** |
| **Degree (sd)** | 14.51 (2.75) | 9.20 (2.16) | 5.87 (1.77) | 3.70 (1.04) | 6.67 (1.44) | 3.68 (0.80) | 8.85 (1.54) | 6.17 (1.19) |
| **Global efficiency (sd)** | 0.42 (0.02) | 0.34 (0.05) | 0.28 (0.06) | 0.16 (0.08) | 0.36 (0.05) | 0.20 (0.08) | 0.34 (0.01) | 0.25 (0.05) |
| **Density (sd)** | 0.08 (0.01) | 0.06 (0.01) | 0.05 (0.01) | 0.04 (0.00) | 0.09 (0.02) | 0.09 (0.01) | 0.05 (0.00) | 0.05 (0.01) |
| **Transitivity (sd)** | 0.37 (0.02) | 0.37 (0.02) | 0.40 (0.05) | 0.45 (0.05) | 0.53 (0.05) | 0.63 (0.08) | NA | NA |
| **Assortativity (sd)** | | | | | | | | |
| ***By degree*** | -0.10 (0.09) | -0.20 (0.09) | 0.22 (0.10) | 0.21 (0.12) | 0.13 (0.10) | 0.19 (0.22) | -0.36 (0.12) | -0.56 (0.08) |
| ***By ward*** | 0.55 (0.04) | 0.68 (0.04) | 0.73 (0.10) | 0.87 (0.06) | 0.68 (0.05) | 0.83 (0.06) | 0.42 (0.06) | 0.57 (0.07) |

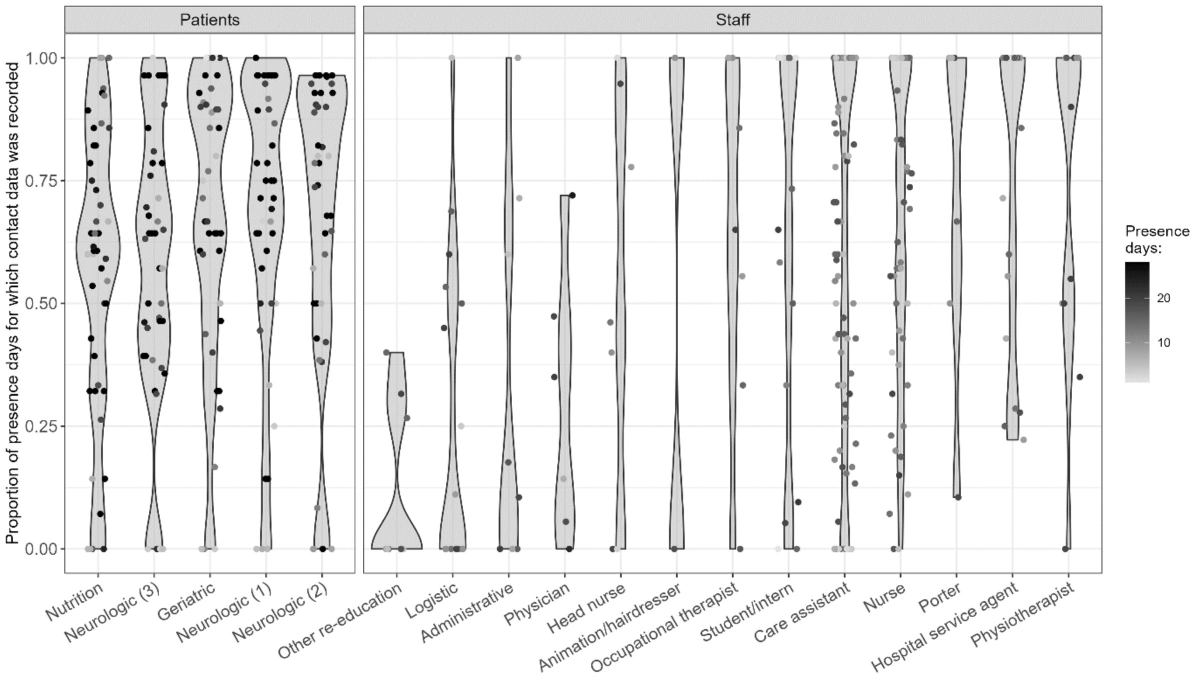


**Supplementary Figure 1: Proportion of presence days during which contact data were recorded, by patient ward and staff category.** Each point is one individual's proportion, calculated over the entire study period. The shading of the points indicate the number of presence days for each individual (darker = more presence days).

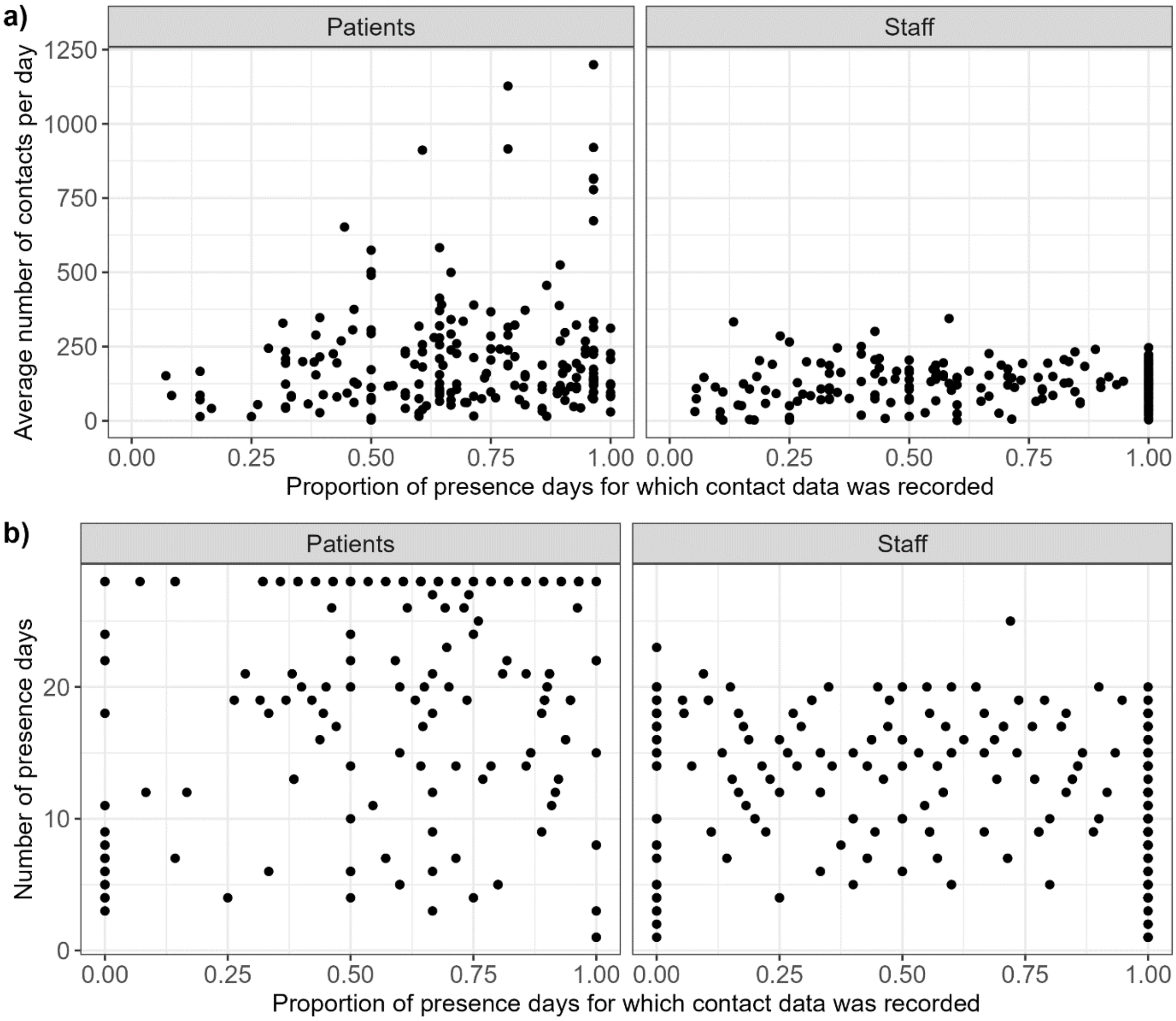


**Supplementary Figure 2: Proportion of presence days during which contact data were recorded is not correlated with a) average number of contacts per day nor b) number of presence days.** Each point is one individual.

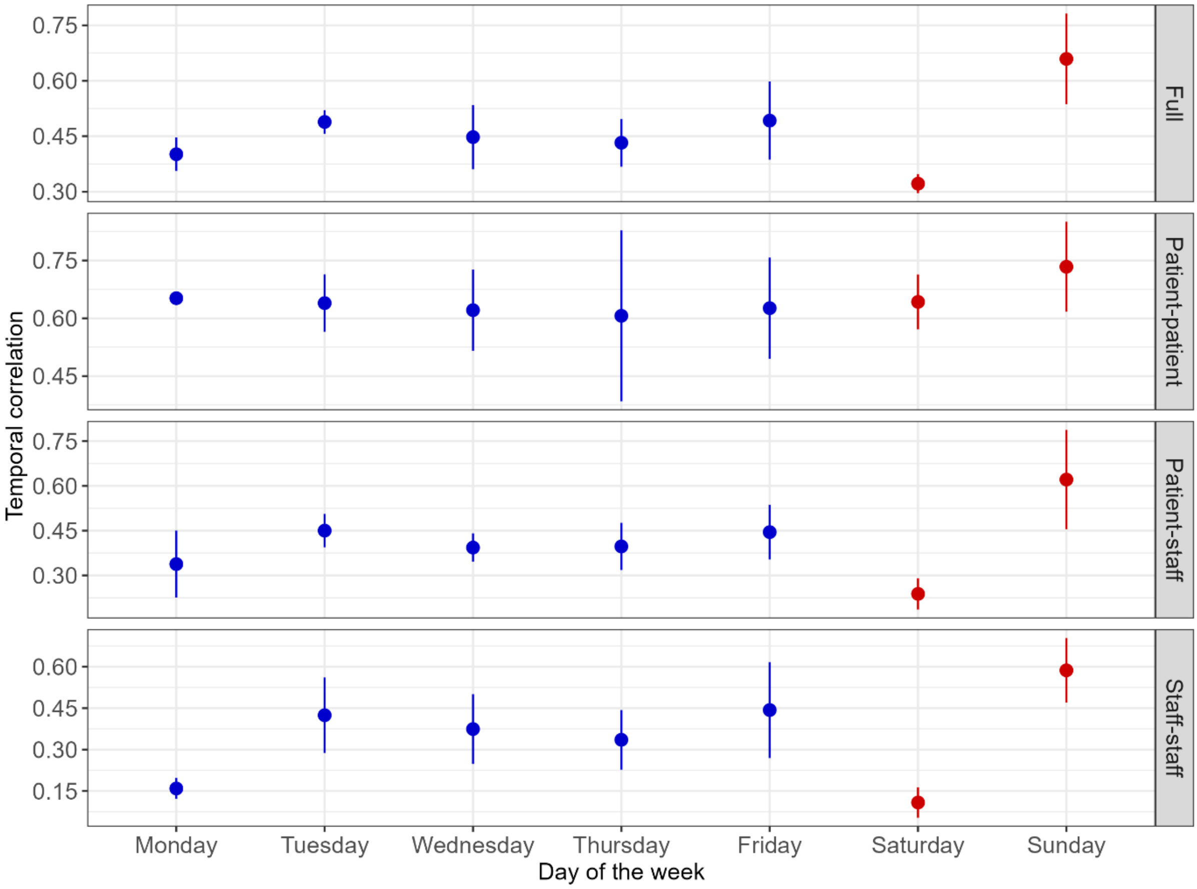


**Supplementary Figure 3: Temporal correlation by subgraph and day of the week.** The correlation is calculated for each day by comparing it to the previous day; for example, the value on "Monday" indicates the correlation between the network on Monday and Sunday. Points indicate the mean correlation, and lines indicate the 95% confidence interval (1.96 times the standard deviation). Weekdays are shown in blue, and weekends in red.

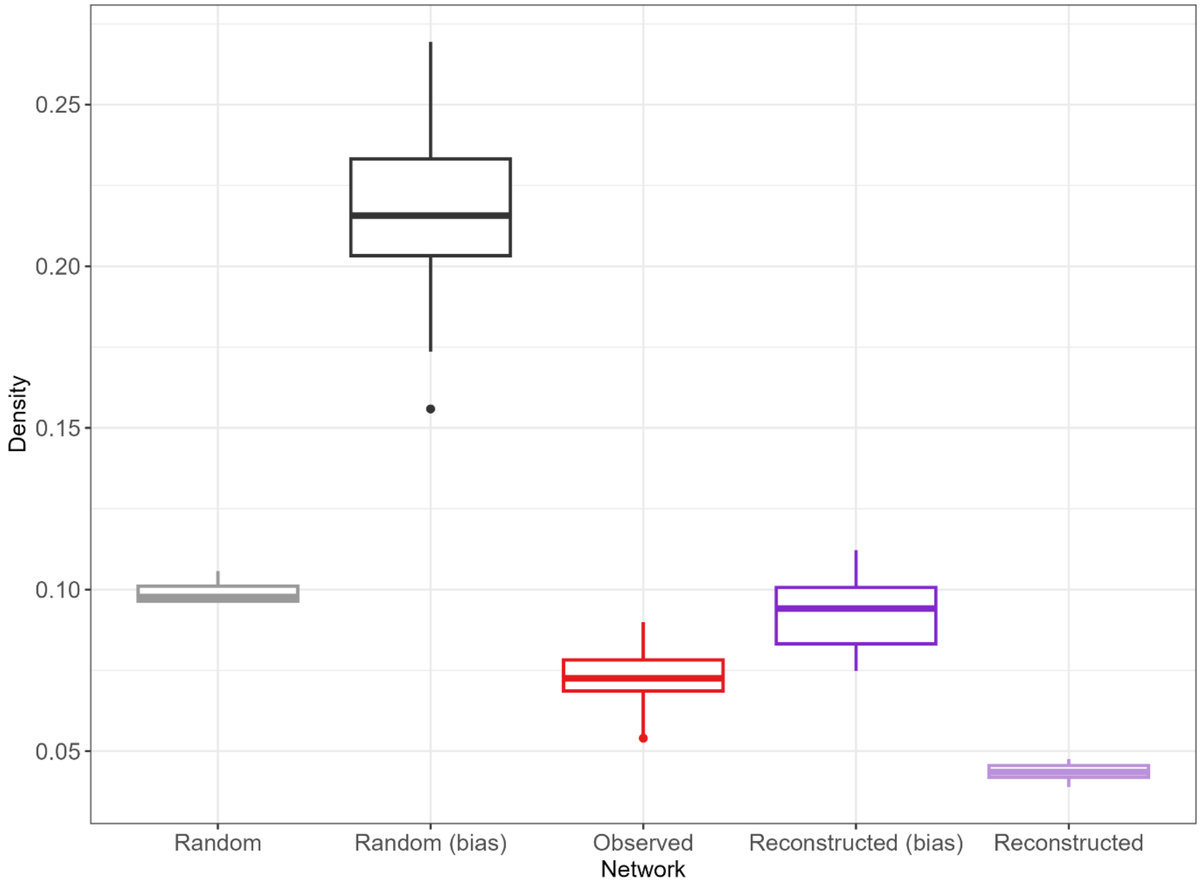


**Supplementary Figure 4: Comparison of density across networks.** The reconstructed networks with observation bias exclude individuals from the network at times when they were known to not wear their sensors. The random networks did not take into account the ward-level structure of the contacts or the probability of recurring contacts. Boxplots for the observed network show the distribution of values calculated for each day. Boxplots for all reconstructed and random networks show the distribution of the median values calculated for each day across 100 networks.

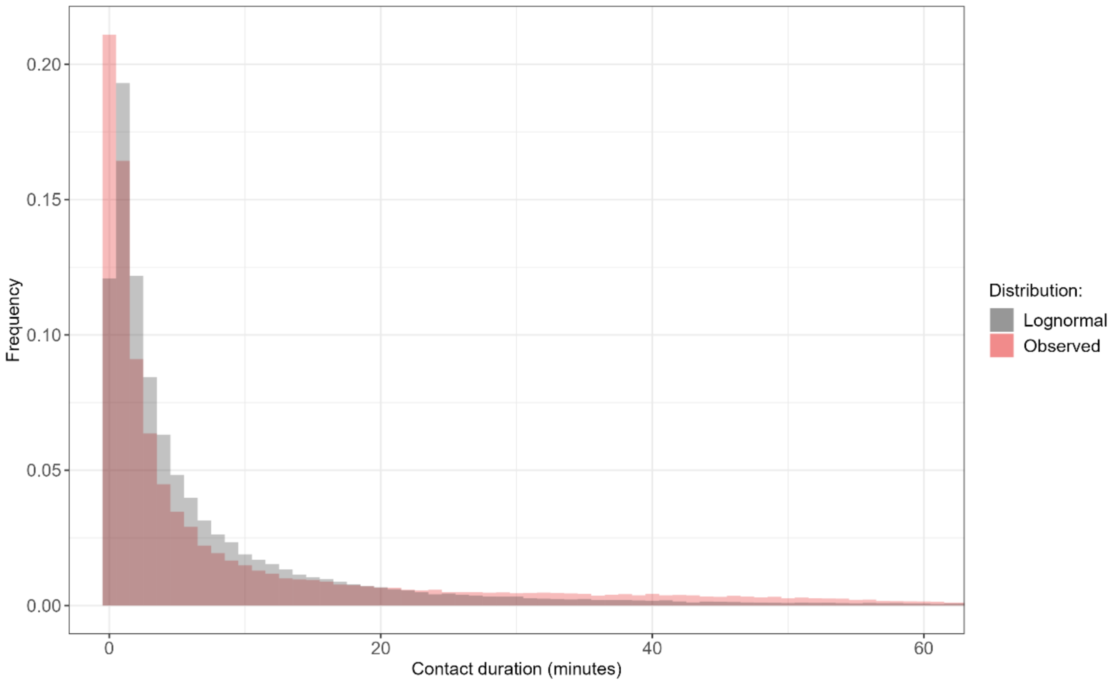


**Supplementary Figure 5: Contact rates in the i-Bird data and sampled from a lognormal distribution.** 100,000 samples are taken from a lognormal distribution is informed by the mean and variance estimated from the data. For ease of visualisation, the x-axis is truncated at 60 minutes.

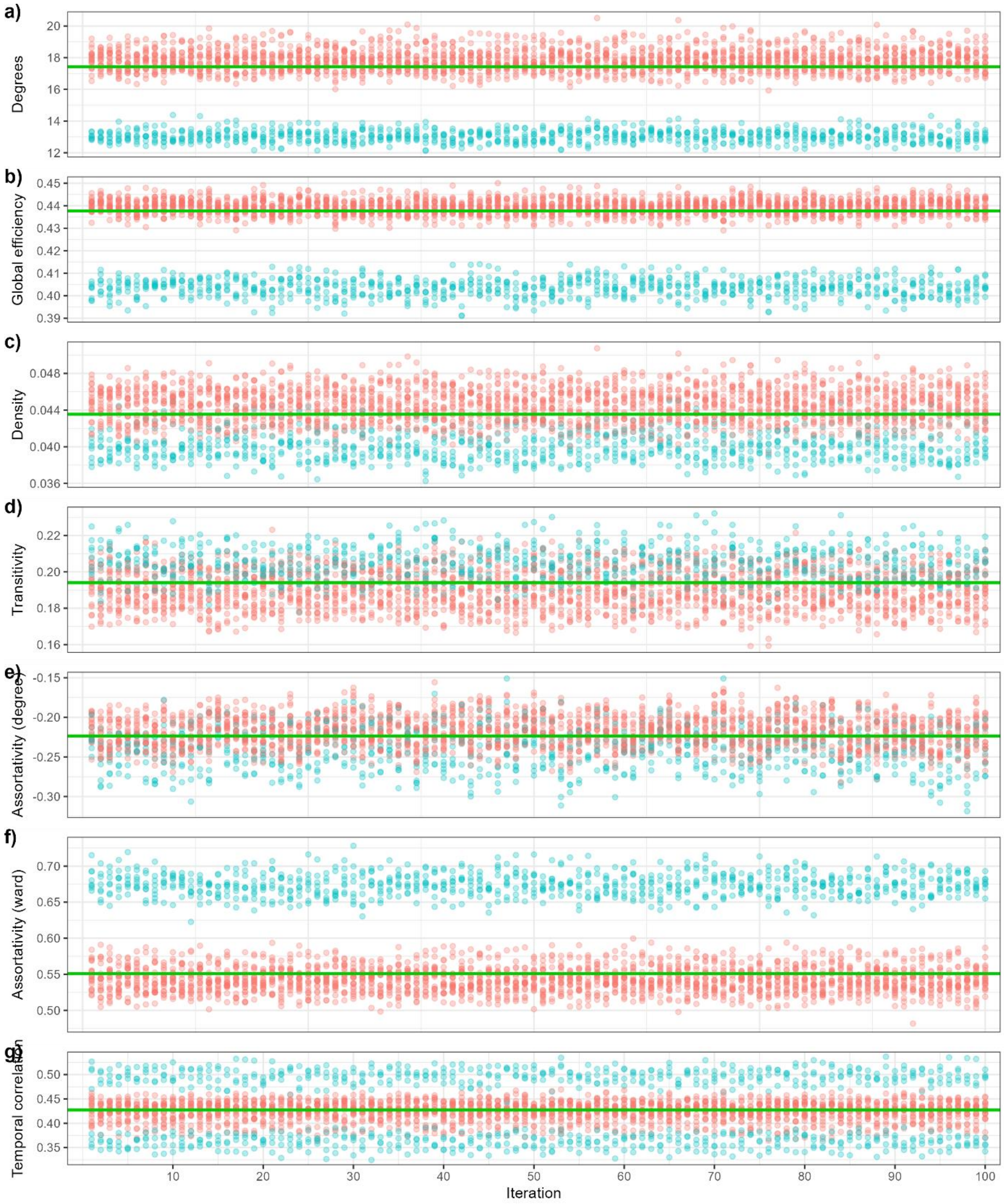


**Supplementary Figure 6: Variability of network characteristics across iterations.** Here, 100 reconstructed networks without bias were generated independently, all informed by the same contact rates estimated from the i-Bird data. For each iteration, the distribution of the metric calculated for each day is shown. Red points are weekdays, and blue points are weekends. The green lines indicate the median value for the correspond metric across all networks and days. Only the distributions of assortativity by degree are significantly different between networks (Kruskal-Wallis test, p value < 0.001).

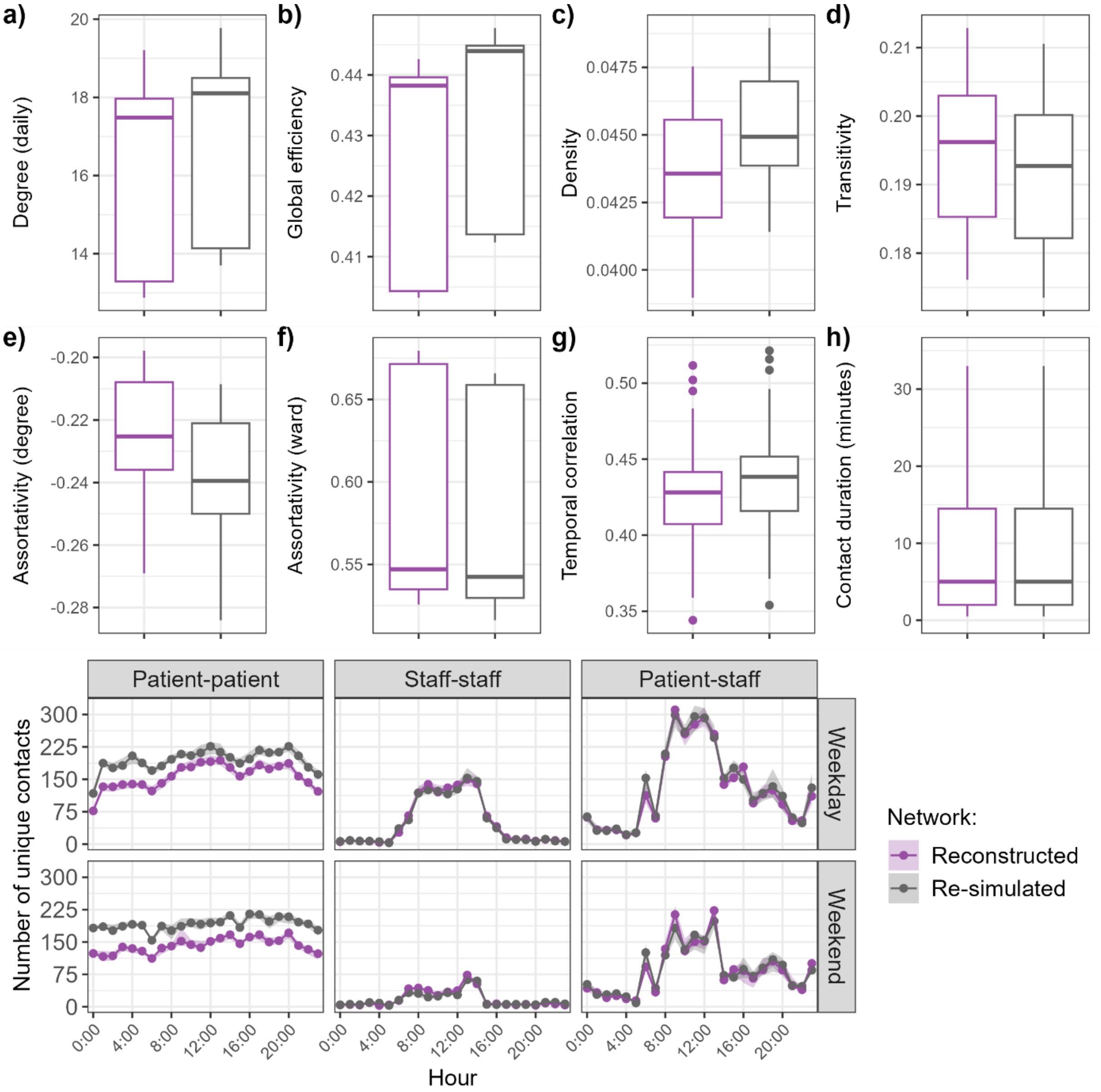


**Supplementary Figure 7: Full reconstructed networks informed by the observed network, compared to full re-simulated networks informed by the reconstructed network with observation bias.**

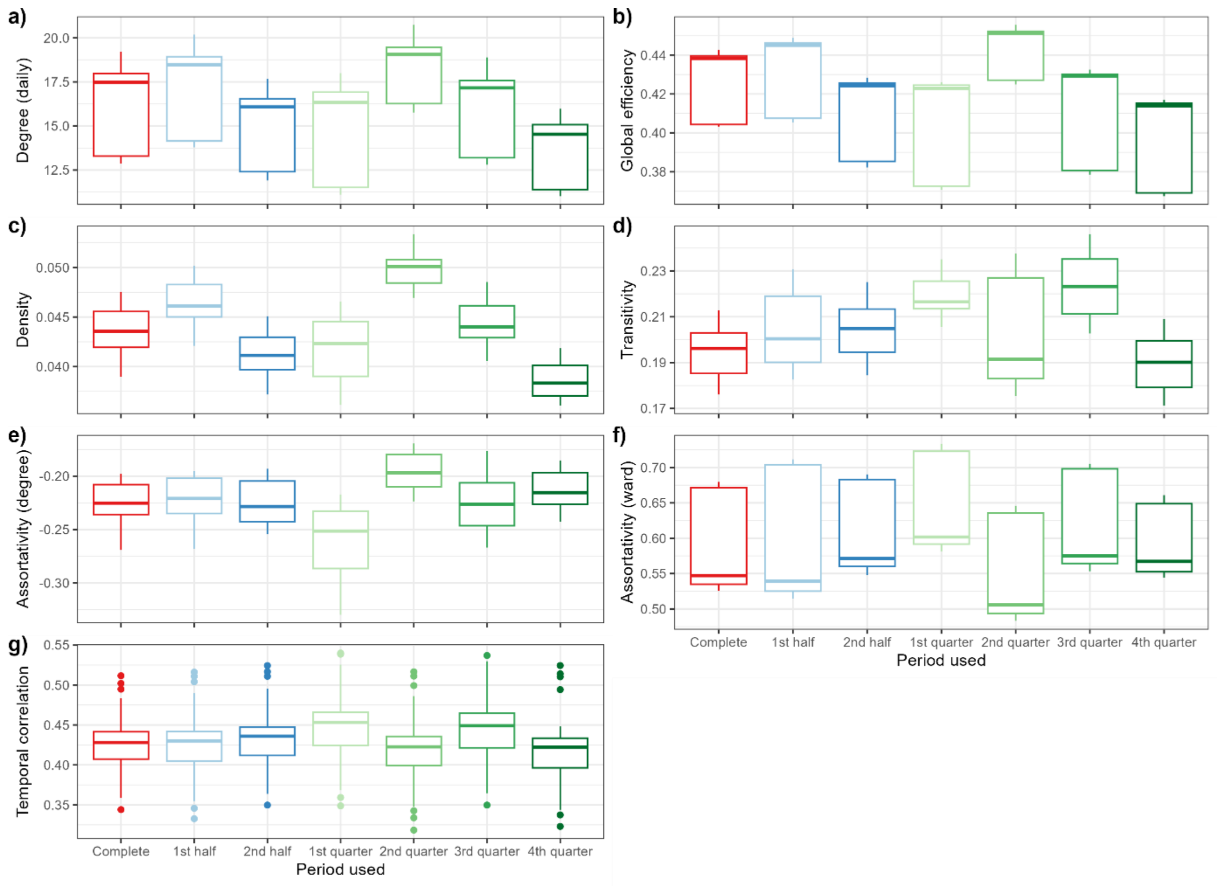


**Supplementary Figure 8: Characteristics for the full reconstructed network, estimated using two weeks or one week instead of the complete observed four weeks.** 1st quarter: 26/07 – 02/08, 2nd quarter: 03/08 – 09/08, 3rd quarter: 10/08 – 16/08, 4th quarter: 17/08 – 23/08. "1st half" includes the 1st and 2nd quarters, and "2nd half" includes the 3rd and 4th quarters.

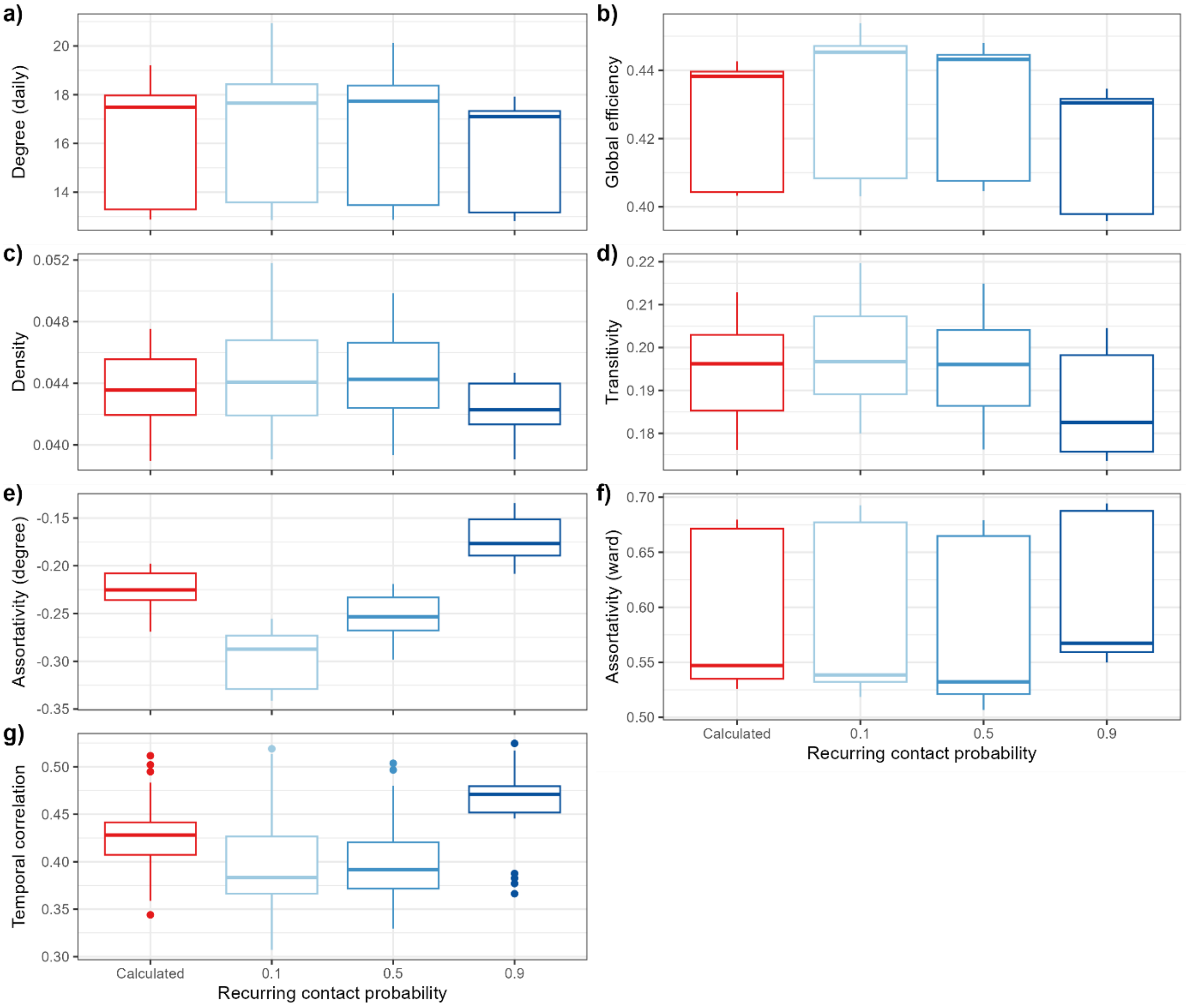


**Supplementary Figure 9: Characteristics for the full reconstructed network when manually setting recurring contact probability to 0.1, 0.5 or 0.9.** The calculated probabilities are 0.78 for patients and 0.71 for staff. In the other scenarios, the probabilities for patients and staff are set to the same value.
